# Genome-wide association study in 404,302 individuals identifies 7 significant loci for reaction time variability

**DOI:** 10.1101/2023.04.03.23288056

**Authors:** Olivia Wootton, Alexey A. Shadrin, Christine Mohn, Ezra Susser, Raj Ramesar, Ruben C. Gur, Ole A. Andreassen, Dan J. Stein, Shareefa Dalvie

## Abstract

Reaction time variability (RTV), reflecting fluctuations in response time on cognitive tasks, has been proposed as an endophenotype for many neuropsychiatric disorders. There have been no large-scale genome wide association studies (GWAS) of RTV and little is known about its genetic underpinnings. Here, we used data from the UK Biobank to conduct a GWAS of RTV in participants of white British ancestry (*n* = 404,302) as well as a trans-ancestry GWAS meta-analysis (*n* = 44,873) to assess replication. We found 161 genome-wide significant single nucleotide polymorphisms (SNPs) distributed across 7 genomic loci in our discovery GWAS. Functional annotation of the variants implicated genes involved in synaptic function and neural development. The SNP-based heritability (*h*^*2*^_SNP_) estimate for RTV was 3%. We investigated genetic correlations between RTV and selected neuropsychological traits using linkage disequilibrium score regression, and found significant correlations with several traits, including a positive correlation with schizophrenia. We assessed the predictive ability of a polygenic score (PGS) for RTV, calculated using PRSice and PRS-CS, and found that the RTV-PGS significantly predicted RTV in independent cohorts, but that the generalizability to other ancestry groups was poor. These results identify genetic underpinnings of RTV, and support the use of RTV as an endophenotype for neurological and psychiatric disorders.

## Introduction

Elevated intra-individual variability in reaction time, namely increased trial-by-trial fluctuations in response time on cognitive tasks, has been associated with neurodevelopmental and neurodegenerative disorders.^1^ Increased variability in reaction time is thought to reflect disruptions in attentional control and executive function and it has been associated with abnormalities in brain structure and function.^1-4^ Increased reaction time variability (RTV) has been demonstrated in attention-deficit hyperactivity disorder (ADHD),^5,6^ schizophrenia,^4,7^ bipolar disorder,^8^ and major neurocognitive disorders.^9^ The heritability of RTV has been established in twin and family studies (h^*2*^ = 0.28 – 0.5)^10,11^ and consequently, RTV has been proposed as an endophenotype for some of these disorders.

Despite increasing interest in the genetic basis of RTV, its genetic architecture remains poorly understood. A limited number of candidate gene studies have provided evidence for an association of RTV with catecholamine system genes.^12-14^ There has only been one genome-wide association study (GWAS) of RTV to date (*n* = 857), which identified one genome-wide SNP, rs62182100.^15^ The significant SNP is an intronic variant located within the *HDAC4* gene, which plays a role in transcriptional regulation and has been implicated in synaptic plasticity, learning and memory.^16^ However, due to the small sample size and the lack of independent replication in this GWAS, insight into the genetic underpinnings of reaction time variability remains limited. A GWAS with a larger sample size may facilitate the identification of more significant loci and provide the power needed for a more comprehensive investigation of the genetic architecture of this trait.

Here, we conduct the largest GWAS of RTV to date with a sample size of 404,302 individuals using data from the UK Biobank. We aim to identify common genetic variants and genes associated with RTV, and we calculate the first SNP-based heritability (*h*^*2*^_SNP_) estimate for the trait. We also calculate estimates of genetic correlations with neurodevelopmental disorders and other phenotypes that have been previously associated with RTV. Lastly, we test the external validity of our results by performing polygenic prediction of RTV in independent samples of European and African ancestries.

## Materials and Methods

### Participants and Phenotype Definition

This study used data from the UK Biobank (UKB), obtained under accession number 27412. The UKB is a large-scale biomedical database with genotype and phenotype data for approximately 500,000 individuals.^17^ At baseline, a brief cognitive assessment, including a custom-made reaction time test, was administered to participants (aged 40 – 70 years) as part of the fully-automated questionnaire. The UKB reaction time test is based on a Go/NoGo test and is designed to measure processing speed.^18^ Participants were shown 2 cards with symbols on them and asked to push a button as quickly as possible when the symbols on the card matched. The test consisted of 12 trials, 9 of which contained matching cards. The UKB reaction time test has demonstrated good concurrent validity with well-validated tests of reaction time.^18^ RTV was operationalized as the intra-individual standard deviation (ISD) of reaction times across correct trials. Prior to calculating the ISD, trials with a reaction time < 50ms (suggesting anticipation instead of reaction), and > 200ms (indicating a response after the cards had disappeared) were excluded. ISD scores were calculated for participants with ≥ 3 correct trials. As RTV was non-normally distributed, RTV values were rank-based inverse normal transformed. For the discovery dataset, we included 405,022 individuals with “white British” ancestry (54% females; mean age 56.88 years), classified according to self-declared ethnicity and genetic principal component analysis. We used all other ancestry groups from the UKB for replication analysis - this included participants who completed the UKB reaction time test with a self-reported ethnicity of “white non-British” (*n* = 28,600), “Asian or Asian British” (*n* = 8,904), or “Black or Black British” (*n* = 7,415), totaling to 44,919 individuals (55% female, mean age 54.27 years) for inclusion in the replication GWAS.

### Genome-wide association analysis

GWAS was conducted using version 3 of the UKB genetic data. Genotyping, imputation, and central quality control procedures for the UKB genotypes are described in detail elsewhere.^19^ The REGENIE method was used and involves 2 steps. In step 1, polygenic predictors are constructed by fitting a whole genome regression model to the UKB genotype data. Additional quality control filters were applied to the UKB genotype calls using PLINK 2.0^20^ prior to conducting step 1 of REGENIE. Quality control steps included removing: 1) individuals with > 10% missing genotype data, 2) SNPs with >10% genotype missingness, 3) SNPs failing the Hard-Weinberg equilibrium tests at *p* = 1 × 10^−15^, and 4) SNPs with a minor allele frequency (MAF) < 1% or minor allele count (MAC) < 50. After quality control, 582,052 variants and 405,019 samples were included in step 1 of REGENIE. In step 2 of REGENIE, a linear regression model was used to test for phenotype-genotype associations using imputed UKB genotype data, conditional upon the predictions of the model from step 1. The association model in step 2 included age, sex and the first 10 genetic principal components as covariates. Variants with an INFO score < 0.8 and MAC < 20 were excluded in step 2 leaving 19,963,755 SNPs and 404,302 samples for inclusion in the GWAS.

### Replication cohort and meta-analysis

We sought to replicate the lead SNPs from the discovery GWAS in an independent association analysis. First, we used REGENIE to conduct association analysis within all other ancestry groups (“white non-British”, ““Asian or Asian British”, and “Black or Black British”) from the UKB separately. Quality control procedures were identical to those used for the discovery analysis. Following GWAS, the summary statistics for 28,396,731 SNPs (*n* = 44,873 after quality control) were meta-analysed using an inverse variance based approach implemented in METAL^21^. To assess for replication, we determined whether lead SNPs from the discovery GWAS reached significance in the replication GWAS (α = 0.05/7; *p* < 0.0071). Additionally, we examined if the effect directions of the A1 allele of lead SNPs from the discovery GWAS were concordant across the discovery and replication GWAS. A binomial test was performed using R v4.1.0^22^ to assess for an excess or deficit of concordant SNPs than would be expected by chance. Lastly, we used METAL to conduct an inverse variance-weighted meta-analysis of the discovery and replication GWAS.

### Genomic risk loci characterization

Genomic risk loci for RTV were characterized from the GWAS results using Functional Mapping and Annotation of Genome-Wide Association Studies (FUMA)^23^. First, the SNP2GENE function was used to identify independent significant SNPs, defined as SNPs with a *p*-value ≤ 5 × 10^−8^ and independent of other genome wide significant SNPs at r^2^ < 0.6. The correlation estimates were calculated using the 1000 Genomes Project Phase 3 release European reference panel^24^. A genomic risk locus included all SNPs, including those from the reference panel, that were in linkage disequilibrium of r^2^ ≥ 0.6 with an independent significant SNP. Genomic risk loci that were within 250 kilobases (kb) of each other were merged into one locus. Lead SNPs were defined as independent significant SNPs that were independent of each other at r^2^ < 0.1. Regional visualization plots were produced using LocusZoom^25^.

### Functional mapping and annotation

The independent significant SNPs and SNPs in LD (r^2^ > 0.6) with the independent significant SNPs (henceforth referred to as candidate SNPs) were functionally annotated using ANNOVAR^26^, combined dependent depletion (CADD)^27^, RegulomeDB (RDB)^28^, and 15-core chromatin states^29^. The NHGRI-EBI GWAS catalogue was searched to assess for previous associations of the candidate SNPs. eQTL mapping for significant SNP-gene pairs (FDR *q* < 0.05) was performed using GTEx v8 whole blood and brain tissue (http://www.gtexportal.org/home/datasets), RNAseq data from the CommonMind Consortium^30^, and the BRAINEAC database (http://www.braineac.org/).

Identified lead SNPs were mapped to likely target genes using The OpenTargets Variant-to-Gene pipeline which integrates a positional score (based on distance to the canonical transcription start site) with data from quantitative trait loci and chromatin interaction experiments and *in silico* functional predictions.^31,32^ For each lead SNP, we also report the nearest gene identified through positional mapping using FUMA. Gene-based analysis of 19,129 protein coding genes was performed using MAGMA^33^ as implemented in FUMA, with a SNP-wise mean model and the 1000 genomes project phase 3 release European reference panel. To control for multiple testing, a Bonferroni corrected *p*-value was used (α = 0.05/19,129 genes tested; *p* < 2.61 × 10^−6^). Additionally, gene-set enrichment analysis was conducted using: 1) significant genes from MAGMA gene-based analysis, 2) genes identified through the OpentTargets’ Variant-to-Gene pipeline, and 3) genes identified through positional mapping in FUMA. Hypergeometric tests were applied through the GENE2FUNC function in FUMA to assess if the identified genes were over-represented in 15,496 gene sets obtained from MsigDB v7.0^34^. Bonferroni correction for multiple testing was applied and gene sets with *p* < 3.23 × 10^−6^ were considered significant.

### Heritability, polygenicity, and discoverability

We used univariate GCTA-GREML analysis^35^ and MiXeR^36^ to estimate the proportion of variance explained by common genetic factors, i.e. *h*^*2*^_SNP_. The covariates included in the GCTA-GREML analysis were the same as those included in the GWAS. The proportion of causal variants (polygenicity) and the average explained variance per causal variant (discoverability) were estimated using MiXeR v1.2^36^. The univariate mixture model considers MAF, sample size, LD structure and genomic inflation to derive estimates of heritability, polygenicity, and discoverability using maximum likelihood estimation.

### Genetic correlation and phenotypic associations

Genetic correlations between RTV and phenotypes known to be associated with RTV were calculated using linkage disequilibrium score regression (LDSC)^37,38^. Summary statistics for general cognitive ability (GCA), educational attainment, Alzheimer’s disease, post-traumatic stress disorder (PTSD), ADHD, schizophrenia, neuroticism, intracranial volume, cortical surface area, cortical thickness, and 7 subcortical brain volumes (nucleus accumbens, amygdala, brainstem, caudate nucleus, pallidum, putamen, and thalamus) were used to calculate genetic correlation estimates. Supplementary Table 1 provides further details on the sources of the GWAS summary statistics. Using data from the UKB, the relationships between RTV and the same phenotypes as listed above were assessed using linear regression (Supplementary Note; Supplementary Table 2).

**Table 1.** Genome-wide significant loci for the discovery GWAS of RTV in 404,302 individuals

| Genomic Risk Locus | Lead Variant | Chr | Position | A1/A2 | Discovery GWAS |  |  |  |  | Replication GWAS |  |
| --- | --- | --- | --- | --- | --- | --- | --- | --- | --- | --- | --- |
|  |  |  |  |  | A1 Freq | Beta | P-value | Top Assigned Gene | Nearest Gene | Beta | P-value |
| 1 | rs183204237 | 2 | 162188514 | C/G | $9.79 \times 10^{-4}$ | -0.21 | $2.34 \times 10^{-8}$ | *PSMD14* | *PSMD14* | -0.088 | 0.46 |
| 2 | rs35206686 | 4 | 152762934 | T/TC | 0.736 | -0.015 | $6.09 \times 10^{-9}$ | *FHIP1A* | *RP11-424M21* | -0.004 | 0.615 |
| 3 | rs17167210 | 7 | 133339343 | A/G | 0.435 | 0.013 | $1.5 \times 10^{-9}$ | *EXOC4* | *EXOC4* | -0.01 | 0.141 |
| 4 | rs35859241 | 15 | 51736660 | G/A | 0.254 | -0.014 | $3.69 \times 10^{-8}$ | *SCG3* | *RP11-707P17.1* | -0.001 | 0.864 |
| 5 | rs912243472 | 15 | 74096786 | G/GA | 0.135 | 0.018 | $1.97 \times 10^{-8}$ | *TBC1D21* | *INSYN1* | 0.005 | 0.575 |
| 6 | rs1863115 | 17 | 44625928 | A/C | 0.74 | 0.017 | $2.47 \times 10^{-10}$ | *LRRC37A2* | *LRRC37A2* | 0.003 | 0.755 |
| 7 | rs11697176 | 20 | 3831629 | T/C | 0.112 | -0.021 | $1.9 \times 10^{-8}$ | *PANK2* | *MAVS* | 0.008 | 0.58 |

### Polygenic score validation

For polygenic score validation we used controls from two independent cohorts of European and African ancestry, The *Thematically Organised Psychosis (TOP) Study*^39^ and *The Genomics of Schizophrenia in the South African Xhosa People (SAX) Study*^40^, respectively. RTV on a continuous performance test was calculated for 182 healthy controls from the TOP study and 563 controls (people without psychotic disorders) from the SAX study. Additional information on the TOP and SAX study can be found in Supplementary Note 1. We also assessed the predictive ability of a RTV polygenic score (PGS) for RTV in the “white non-British”, “Asian or Asian British”, and “Black or Black British” ancestry groups from the UKB.

The RTV-PGS were calculated from Z-score effect size estimates from the discovery RTV-GWAS using a pruning and thresholding approach implemented in PRSice^41^. Prior to PGS calculation, SNPs with MAF < 0.05 were excluded and pruning was performed using an r^2^ < 0.1 within a 250 kb window. We calculated PGS across 10 p-value thresholds (1, 0.1, 0.05, 0.01, 1 ×10^−3^, 1 × 10^−4^, 1 ×10^−5^, 1 ×10^−6^, 1 ×10^−7^, 5 ×10^−8^) in the white non-British participants from the UKB and linear regression models were used to test the association between RTV and PGS at each threshold. The best performing PGS was used to determine the *p*-value threshold for PGS calculation in all other ancestry groups. Sex, age and the first ten principal components were included covariates. For comparison, we calculated a RTV-PGS in each target cohort using PRS-CS^42^, which uses a Bayesian regression framework and places a continuous shrinkage prior on SNP effect sizes. The 1000 Genomes Phase 3 release European sample^24^ was used as the LD reference panel for PRS-CS. The Bonferroni correction was applied to account for multiple testing (α = 0.05/19 polygenic scores; *p* < 2.63 × 10^−3^).

## Results

### Genome-wide associations

Genome-wide association tests for RTV in the discovery analysis identified 161 genome-wide significant SNPs (*p* < 5 × 10^−8^) (Figure 1; Supplementary Table 3). There were 13 independent significant SNPs distributed across 7 genomic loci (Table 1). Regional visualization plots for the significant loci are depicted in Figure 2 and Supplementary Figure 3. Four of the seven genome-wide significant loci have been reported as significant in previous GWAS of general cognitive ability and intelligence (Supplementary Table 4). None of the risk loci were significant in the previous GWAS of RTV.^15^ The linkage disequilibrium score regression intercept was 1 (SE = 0.01), consistent with minimal inflation of the test statistic due to population stratification.

**Figure 1.**
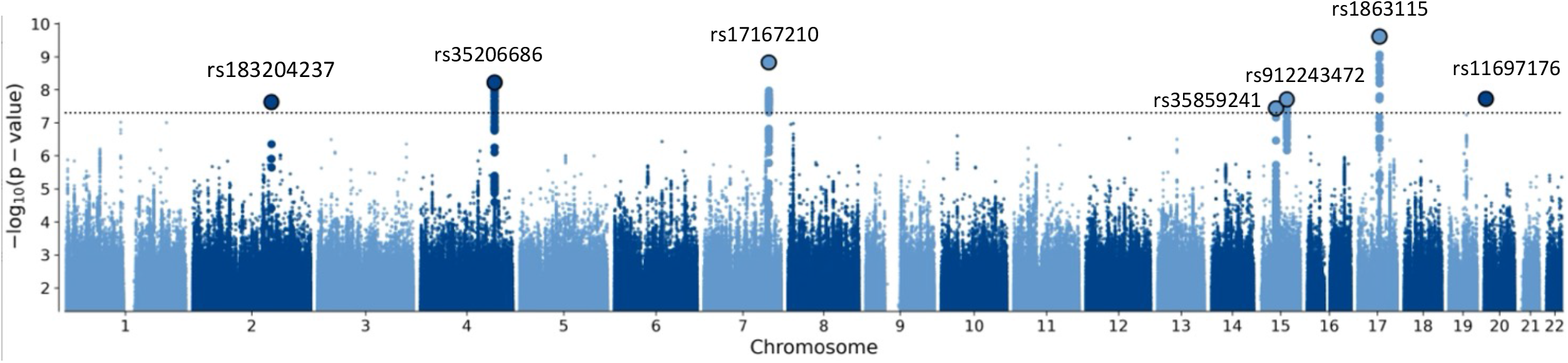
Manhattan plot of discovery GWAS for RTV in the UK Biobank. Manhattan plot for the observed -log10 *p*-values for an association with RTV in the discovery GWAS. The dotted line indicates a genome-wide significance threshold of 5 × 10^−8^. The lead SNPs from the GWAS are outlined in black and the candidate SNPs are shown in bold.

**Figure 2.**
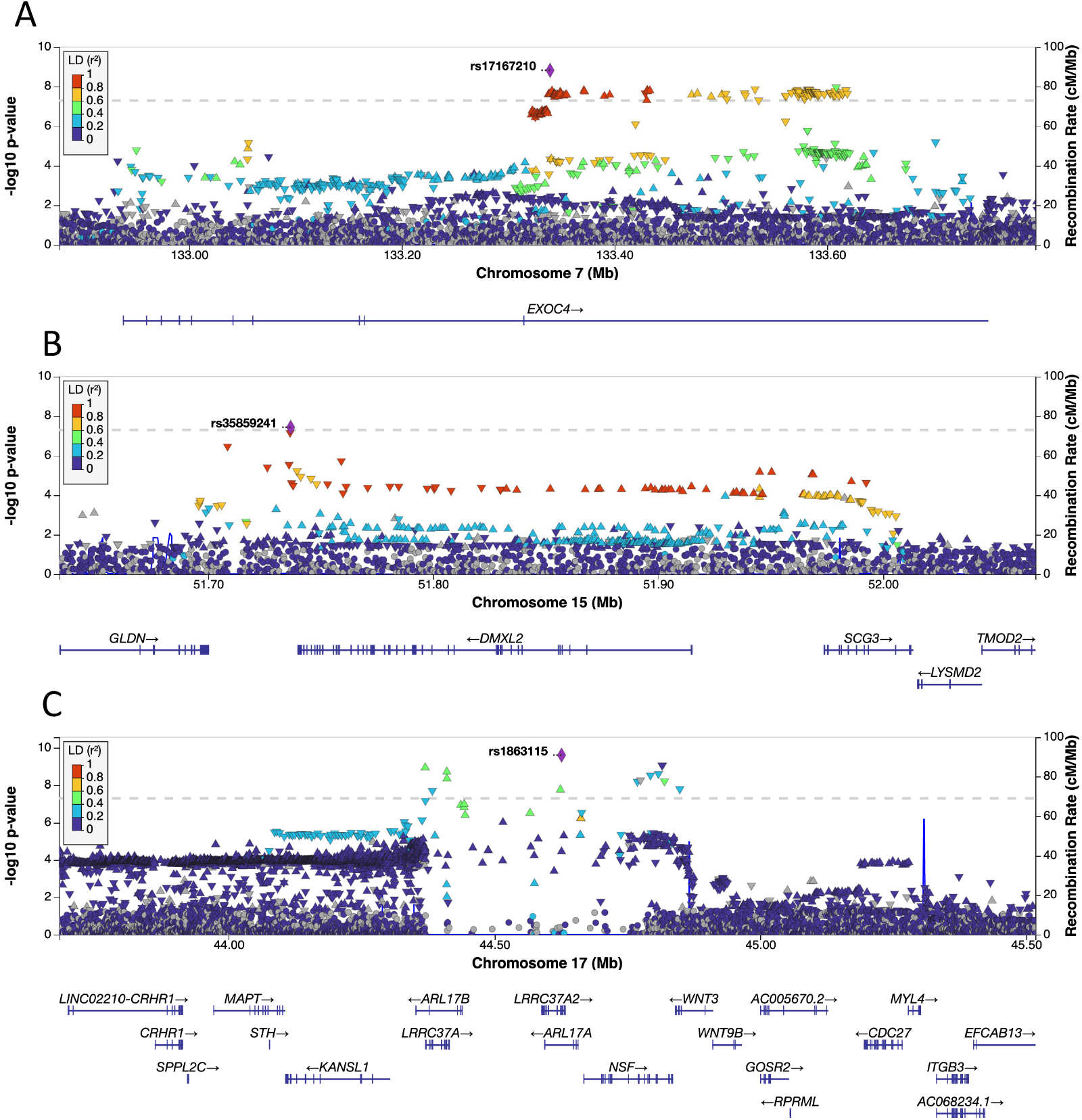
Regional association plots for three genome-wide significant loci in the discovery RTV-GWAS. Regional plots for rs17167210 (**A**), rs35859241 (**B)** and rs1863115 (**C**). The dotted line denotes a genome-wide significance threshold of 5 × 10^−8^. SNPs in the genomic risk loci are colour-coded as a function of their linkage disequilibrium r^2^ to the lead SNP in the region.

None of the lead SNPs from the discovery GWAS reached significance in the replication GWAS (Table 1; Supplementary Figure 1). Due to the limited number of lead SNPs at a genome-wide significant threshold, binomial tests for concordance were performed using lead SNPs from the discovery GWAS at a suggestive threshold of *p* of ≤ 5 × 10^−5^. There were 261 lead SNPs at the suggestive level in the discovery GWAS, with 156 of them having concordant direction of effects (binomial test p = 0.047) in the replication GWAS.

In the meta-analysis of the discovery and replication GWAS (*n* = 449,175), there were 41 genome-wide significant SNPs (*p* < 5 × 10^−8^) distributed across 6 genomic loci (Supplementary Figure 1; Supplementary Table 5). Thirty-six of the genome-wide significant SNPs were also significant in the discovery GWAS. Inspection of the quantile-quantile plot for the meta-analysis shows greater test-statistic inflation above the null for moderately significant *p*-values than in the discovery GWAS (Supplementary Figure 2). The linkage disequilibrium score regression intercept for the meta-analysis was 1 (SE = 0.01), suggesting that the inflation of the test statistic reflects true associations with RTV.

### Integration with functional genomic data

Each lead SNP from the discovery GWAS was mapped to one gene using the OpenTargets Variant-to-Gene pipeline, resulting in 7 mapped genes (Table 1). MAGMA gene-based analysis identified 5 genes significantly associated with RTV: *EXOC4* (*p* = 6.3 × 10^−7^), *TBC1D21* (*p* = 2.57 × 10^−6^), *CNTNAP4* (*p* = 2.59 × 10^−7^), *LRRC37A* (*p* = 9.81 × 10^−10^), and *NSF* (*p* = 4.97 × 10^−7^) (Supplementary Table 6). An additional 17 genes were mapped to candidate SNPs from the discovery GWAS using FUMA positional mapping; resulting in a total of 27 input genes (Supplementary Table 7) for gene set enrichment analysis. Gene set enrichment analysis did not identify any significant gene sets associated with RTV.

The lead variant for the GWAS, rs1863115 (*p* = 2.47 × 10^−10^) (Figure 2, C), is a non-synonymous exonic variant for *LRRC37A2* and an intronic variant for *ARL17A*. The CADD score for rs1863115 is 18.32, suggestive of variant deleteriousness. Based on annotation by the OpenTargets genetic platform, the most likely gene affected by this variant is *LRRC37A2*, a gene that encodes an integral component of the cellular membrane. *LRRC37A2* has been associated with intelligence, and mean reaction time in previous GWAS.^43,44^

There was evidence of functionality for variants in genomic risk loci 3 and 4 (Table 1). The lead variant for locus 3, rs17167210 (*p* = 1.5 × 10^−9^), is located in an intron of *EXOC4* and is an eQTL for *EXOC4* and *LRGUK* in brain tissue (CommonMind Consortium) (Figure 2, A). A nearby intronic variant, rs11768150 *(R*^*2*^ *=* 0.88, *p* = 1.71 × 10^−7^), has a CADD score of 13.5, suggestive of variant deleterious, and a RegulomeDB score of 3a, indicating that the variant is likely to be involved in gene regulation. The lead variant for locus 4, rs35859241 (*p* = 3.69 × 10^−8^), is an eQTL for *SCG3* and *GLDN* in brain tissue (CommonMind Consortium, GTEx Brain). This variant is in LD with rs2606134 (*R*^*2*^ = 0.81, *p* = 1.14 × 10^−4^), which is located within the 5’ untranslated region of *SCG3* (Figure 2, B). The SNP, rs2606134, has a CADD score of 13.15 and a RegulomeDB score of 2b, suggesting that this variant may be biologically relevant.

### Estimating heritability, polygenicity, and discoverability

The *h*^2^_SNP_ for RTV was estimated at 0.029 (SE = 0.002) using GCTA-GREML. MiXeR analysis suggested that RTV is highly polygenic with an estimated 6,800 causal variants explaining the *h*^2^_SNP_ for RTV. As expected for a trait with a low *h*^2^_SNP_ and high polygenicity, discoverability was low (σ_2_β = 5.38 × 10^−6^, SD = 2.85 × 10^−7^) indicating that most SNP-associations have a weak effect. Akaike’s Information Criteria (AIC) for MiXeR analysis was 18.39 indicating reliable model fit.

### Genetic correlations and phenotypic associations

We assessed the genetic correlations and phenotypic relationships between RTV and 17 traits that have been posited to be associated with RTV using LDSC and linear regression respectively (Figure 3; Supplementary Table 8 and 9). After Bonferroni correction, we found significant genetic correlations (α = 0.05/17; *p* < 2.9 × 10^−3^) between RTV and general cognitive ability (r_g_ = -0.44, SE = 0.03), educational attainment (r_g_ = -0.23, SE = 0.03), schizophrenia (r_g_ = 0.26, SE = 0.03), and neuroticism (r_g_ = 0.13, SE = 0.028). The analysis of phenotypic data from the UKB revealed a significant relationship between RTV and several traits, including those that showed significant genetic correlations with RTV (Figure 3, Supplementary Table 9).

**Figure 3.**
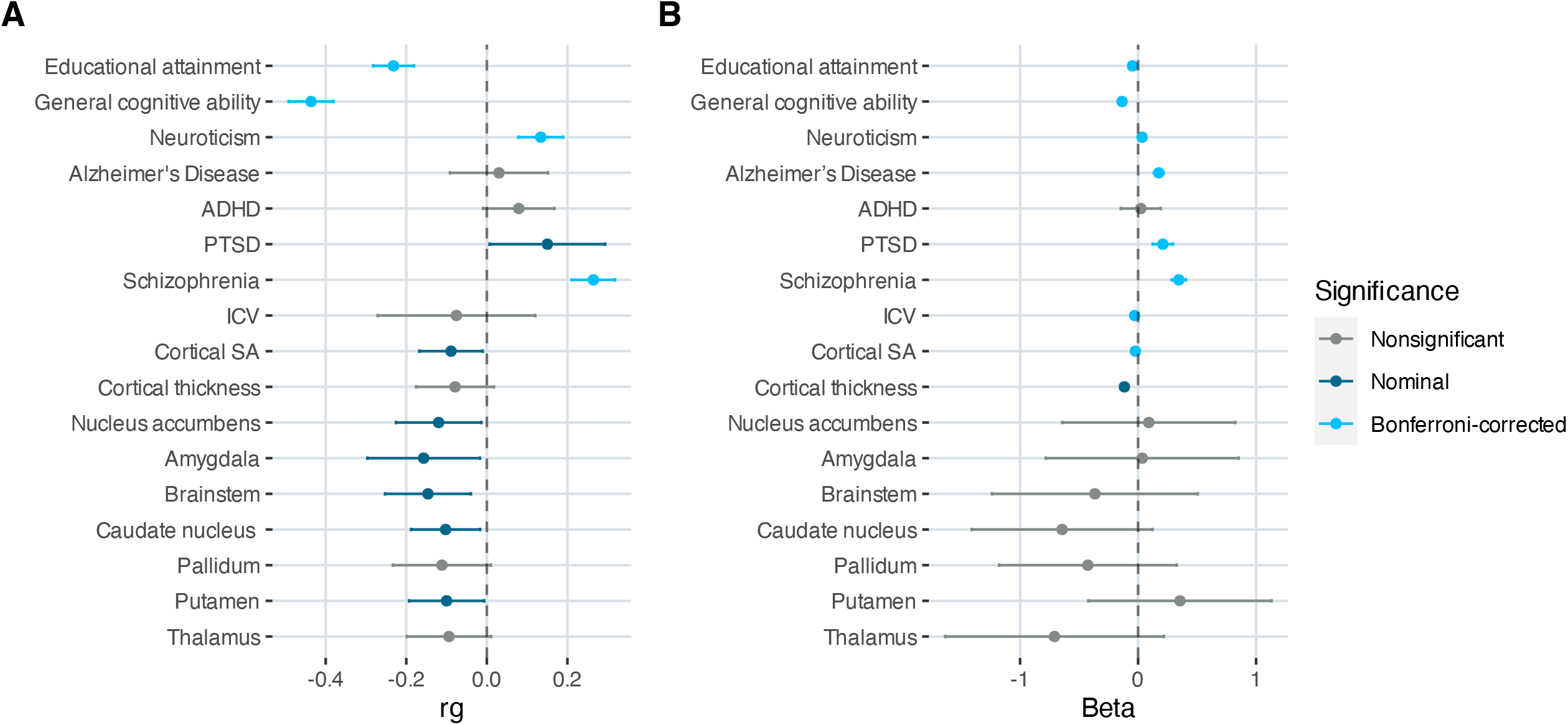
Genetic correlations and phenotypic associations between RTV and 17 selected traits. Genetic correlations were calculated with LD score regression using SNP summary statistics from discovery RTV-GWAS and publicly available summary statistics for other traits (**Supplementary Table 1**). Associations between RTV and the same 17 traits were calculated using phenotypic data from the UK Biobank (**Supplementary Methods)**. Point estimates for correlations and beta coefficients are shown with 95% confidence intervals. Dark blue dots indicate nominally significant *p*-values and light blue dots indicate significant *p*-values after Bonferroni correction.

### Polygenic prediction of RTV

To evaluate the replicability and predictive ability of the results from our discovery GWAS, we calculated a PGS for RTV in the five independent target samples using PRSice and PRS-CS. For the PGS calculated using PRSice, the most significant association between the RTV-PGS and RTV in non-British European participants from the UKB was achieved when all SNPs surviving LD pruning (*n* = 166,662) were included in the PGS calculation (*p*-value threshold = 1) (Supplementary Table 10). The variance explained by this PGS was *r*^2^ = 0.0027 (*p* = 6.09 × 10^−18^). The PRSice RTV-PGS performed poorly in the other ancestry groups from the UKB and there were no significant associations between the PGS and RTV in the South Asian or African ancestry groups (Figure 4). There was an improvement in predictive power when using PRS-CS to calculate the PGS and there was a significant association between the RTV-PGS and RTV in the non-British European (*r*^2^ = 0.0048, *p* = 3.08 × 10^−31^) and South Asian ancestry groups (*r*^2^ = 0.0012, *p* =1.73 × 10^−3^) from the UKB (Figure 4). The PRSice and PRS-CS RTV-PGS did not predict RTV in controls from the TOP and SAX study (Supplementary Table 10; Supplementary Figure 4).

**Figure 4.**
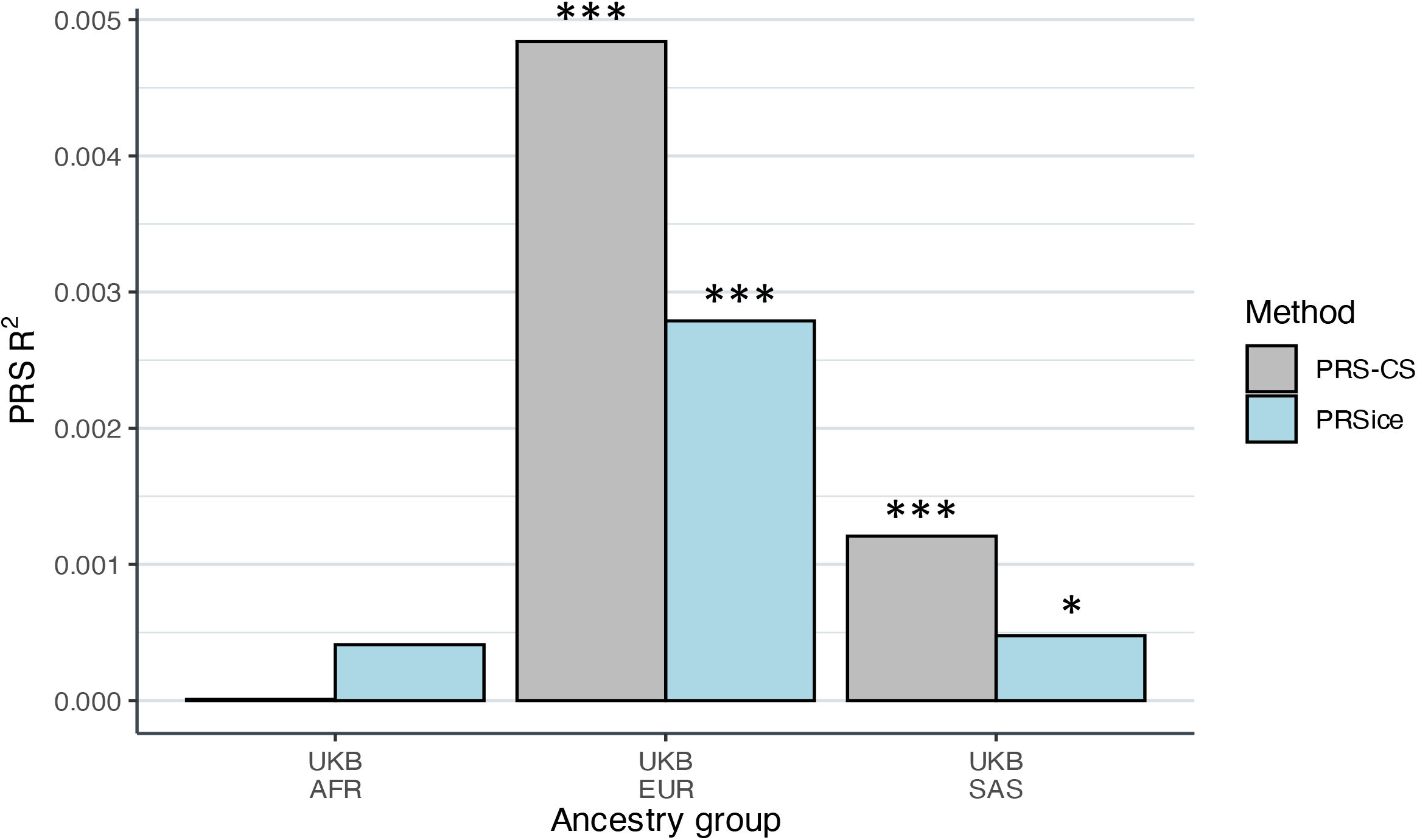
Bar chart showing the predictive accuracy of the RTV-PGS in three independent cohorts. Prediction of RTV by polygenic score (PGS) in the African, non-British European, and South Asian ancestry groups from the UK Biobank. The predictive accuracy of the PGS (R^2^) was assessed in each cohort for a PGS calculated using two methodologies, PRSice and PRS-CS. PRSice PGS were calculated using all single nucleotide polymorphisms surviving LD pruning from the discovery GWAS (*p-value* threshold of 1). \**p*<0.05, \*\*\**p*<2.63 × 10^−3^

## Discussion

Using UKB data, we have performed the largest GWAS of RTV to date and have made several contributions to our understanding of the genetic basis of this cognitive trait. We identified 161 genome-wide significant SNPs for RTV distributed across 7 genomic loci, all of which are novel for RTV. We identified several genes that may play a role in RTV, many of which have been associated with cognitive traits previously. We provide the first SNP-based heritability estimate for RTV, and the first estimates for genetic correlations between RTV and several neuropsychiatric traits. We demonstrate that RTV-PGS derived from the discovery GWAS can significantly predict RTV in an independent cohort, but that the predictive ability declines if the discovery and target populations are of different ancestries.

The genes identified by the GWAS may provide insight into the biological underpinnings of RTV. Although the exact role that many of the identified genes may play in RTV is unclear, several are worthy of further investigation. For example, two of the significant genes, contactin associated protein family member 4 (*CNTNAP4)* and N-ethylmaleimide sensitive factor, vesicle fusing ATPase (*NSF*) encode proteins that play a role in synaptic function. *CNTNAP4* is involved in the synaptic transmission of dopamine and GABA^45^ and *NSF* regulates glutamate receptor binding activity.^46,47^ Alterations in dopaminergic, glutaminergic and GABAergic activity have been associated with RTV^1,48-50^ and thus, further exploration of the association between RTV and *CNTNAP4* and *NSF* may be warranted. Variants in *EXOC4* and *SCG3* showed evidence of regulatory functionality and variant deleteriousness. Both genes are highly expressed in the brain and have been associated with cognitive traits in previous GWAS.^44,51^ *EXOC4* encodes a component of the exocyst complex which plays a role in multiple physiological processes, including neuronal development.^52,53^ *SCG3* encodes a member of the granin family of neuroendocrine secretory proteins and is involved in secretory granule biosynthesis and the storage and transport of neurotransmitters.^54,55^ Another identified gene, inhibitory synaptic factor 1 (*INSYN1)*, is involved in post-synaptic inhibition in the central nervous system^56^ and may be considered for further study. *INSYN1* is a novel association for a cognitive trait but it has been associated with psychiatric disorders, including ADHD,^57^ PTSD,^58^ and Tourette syndrome.^59^ Many of the identified genes play a role in neural development and synaptic functioning, suggesting an important role for these processes in the biology of reaction time variability.

We reported an h^2^_SNP_ of 3%, high polygenicity, and low discoverability for RTV. It is possible that the high polygenicity, despite the relatively low heritability, may be explained by the range of exogenous factors, such as age, sex, handedness, and treatment effects,^5,60,61^ that influence RTV. We hypothesize that a large proportion of the identified 6,800 causal variants may be associated with these exogenous factors and thus, only have indirect and weak effects on RTV.

We found that a PGS, derived from the RTV-GWAS in white British participants from the UKB, was significantly predictive of RTV in the UKB white non-British participants, explaining 0.5% of the variance in the measure, which is expected with a h^2^_SNP_ of 3%.^62^ The predictive accuracy of the PGS was substantially lower in non-European ancestry populations. This is in keeping with prior work on the generalizability of PGS across ancestrally diverse populations with the predictive accuracy of the PGS decreasing as the genetic distance between the discovery and target populations increases.^63^ These results further emphasize the need to increase the representation of ancestrally diverse populations in genomic studies.

We found significant positive genetic correlations between RTV and schizophrenia, and neuroticism. The result for schizophrenia is consistent with previous findings of increased RTV in people with schizophrenia.^4,7^ It is hypothesized that the elevations in RTV reflect cognitive control deficits that occur in the disorder.^4^ The positive genetic correlation between RTV and neuroticism is supported by our phenotype analysis, which demonstrate a positive relationship between the two phenotypes. To our knowledge, the association between these two traits has not been studied and future research is needed to explore the mechanisms that contribute to a relationship between RTV and neuroticism. There were significant negative genetic correlations between educational attainment, and general cognitive ability. This result is in keeping with the negative relationship between these two traits and RTV on the phenotypic level reported in previous literature.^64,65^

There are some limitations to this study. First, the UKB reaction time test is brief and consists of fewer trials than are typically used in simple reaction time tests. This paucity of trials may have reduced the reliability of the measurement thereby affecting our ability to accurately capture RTV for participants, contributing towards the low estimate for h^2^_SNP_. While the associations between RTV and other mental health and cognitive phenotypes in the UKB are in keeping with the associations observed in previous studies of RTV using validated reaction time tests, future studies should consider using a more comprehensive assessment of reaction time. Second, the assessment of reaction time variability differed between the UKB, SAX, and TOP study and heterogeneity in the phenotype may have affected comparisons of RTV among studies. Third, there is a lack of well-powered studies with which to conduct a replication GWAS. The moderate sample size of the replication study and limitations pertaining to the trans-ancestry replicability of risk variants may account for the non-replication of the lead SNPs from our discovery GWAS. Fourth, the low *h*^*2*^_SNP_ of RTV may have affected the accuracy and predictive power of the RTV-PGS. While this low *h*^*2*^_SNP_ limits the potential use of the PGS to predict RTV, we were still able to fulfil the aim of the polygenic score analyses, which was to evaluate the replicability of the results from the discovery GWAS. Lastly, we used self-reported ethnicity as a population descriptor for participants from the UKB. While using the ethnic groups provided by the UKB facilitates comparability with other studies using the same data, future work should consider alternative population descriptors that are better able to capture genetic variation between groups.

In summary, we have conducted the first large-scale GWAS of RTV using 404,302 samples and identified 7 independent associated loci. Several of the implicated genes are involved in neural development and synaptic function and are known to be associated with other cognitive traits. These findings suggest that disruptions to these processes may affect shared biological mechanisms responsible for maintaining the integrity of various aspects of cognitive function. Despite the relatively low SNP-based heritability of RTV observed in our study, it provides evidence that there is a genetic contribution to the trait. Future studies may leverage these findings to improve our understanding of the genetic mechanisms contributing to RTV and gain novel insight into the biological underpinnings of related complex disorders, like schizophrenia.

## Data Availability

All data produced in the present study are available upon reasonable request to the authors and will be deposited in publicly available repositories once published. Sources of the publicly available summary statistics used in this study are described in Supplementary Table 1.

## Acknowledgements

We would like to thank the participants and members of the research teams involved in the UK Biobank, SAX Study and TOP study. We would like to express our gratitude to Torril Ueland and Beathe Haatviet for their support of this work and for facilitating access to the cognitive data from the TOP study. The current study was supported by National Institute of Mental Health (NIMH: Grant number U01MH125053), and The Research Council of Norway (275054). This research has been conducted using data from UK Biobank, a major biomedical database (Project ID number 27412; www.ukbiobank.ac.uk). This work was partly performed on the TSD (Tjeneste for Sensitive Data) facilities, owned by the University of Oslo, operated and developed by the TSD service group at the University of Oslo, IT-Department (USIT). Computations were also performed using facilities provided by the University of Cape Town’s ICTS High Performance Computing team: *hpc.uct.ac.za*. The SAX study was funded by the NIMH (Grant number: 5UO1MH096754). AS was supported by the EEA and Norway grant (#EEA-RO-NO-2018-0573) and the Research Council of Norway (#326813). OA receives funding from the Research Council of Norway (#276082, #223273). DS, RR and SD are supported by the South African Medical Research Council. The content is solely the responsibility of the authors and does not necessarily represent the official views of the funders.

## Conflicts of Interest

The authors declare the following competing interest: OA has received speaker’s honorarium from Lundbeck and is a consultant for Healthlytix. The remaining authors declare no competing interests.

## Notes

### Author Declarations

This work was approved by the University of Cape Town Human Research Ethics Committee (reference number - 734/2021). The UKB has ethical approval (REC reference number - 11/NW/0382) and is overseen by an Independent Ethics and Governance council. The Genomics of Schizophrenia in the South African Xhosa People Study was approved by the University of Cape Town Human Research Ethics Committee (reference number - 049/2013). The Thematically Organised Psychosis Research Study was approved by the Norwegian Scientific Ethical Committee and the Norwegian Data Protection Agency.

