## Supplementary Note for "Genome-wide association study in 404,302 individuals identifies 7 significant loci for reaction time variability"

**Characterizing the genetic architecture of reaction time variability using a genome-wide approach**

### 1. **Supplementary Information**

#### 1.1. **Additional cohort descriptions and methodology**

This study used reaction time data from 2 independent studies for polygenic risk score analyses. The studies and operationalization of RTV in the studies are described in detail below.

##### 1.1.1. **The Genomics of Schizophrenia in the South African Xhosa People Study (SAX**)

The SAX study is a case-control study that aimed to characterize the genetic architecture of schizophrenia in the Xhosa population of South Africa.^1^ The study enrolled 2,092 individuals of African ancestry. Individuals with schizophrenia (cases) were recruited from inpatient psychiatric units and outpatient clinics in the Eastern Cape and Western Cape Provinces of South Africa. Controls were recruited from outpatient clinics and were matched to cases for age, gender, education and region of recruitment. Cases were individuals with a 2 year history of schizophrenia or schizoaffective disorder confirmed using the Structured Diagnostic Interview for DSM-IV Axis I Disorders.^2^ Informed consent was obtained from all participants and the study was approved by the University of Cape Town Human Research Ethics Committee (reference number – 049/2013).

DNA samples were obtained from participants and have been genotyped using the Affymetrix SAXv2 chip (Affymetrix Inc., Santa Clara, CA, USA). Quality control was performed using PLINK v1.9^3^ and included the removal of 1) individuals with > 5% missing data, 2) individuals with heterozygosity rate > 3 standard deviations from the mean, 3) samples with discrepancies between reported and genetically determined sex, 4) related individuals (IBD > 0.2), 5) SNPs with genotype missingness rate > 5%, 2) SNPs that deviated from Hardy-Weinberg equilibrium with p<1x10-10 in cases and p<1x10-6 in controls, 6) SNPs with a minor allele frequency (MAF) < 5%. Data was uploaded to the Wellcome Trust Sanger Institute Imputation Server (<https://imputation.sanger.ac.uk/>)^4^ for phasing and imputation. Phasing was performed using EAGLE2^5^ and imputation by the Positional Burrows Wheel Transform tool^6^. The African Genome Resources haplotype reference panel^7^ was used for imputation. Variants with a posterior genotype probability < 0.9 were set as missing. Post-imputation quality control included the removal of multi-allelic variants and variants with INFO score < 0.7 or MAF < 5%.

An adapted version of the University of Pennsylvania Computerized Neurocognitive Battery (PennCNB)^8^ was completed by a subsample of participants. Reaction time was measured during the Penn Continuous Performance Test (PCPT)*^9^*. During the PCPT, participants are shown 7-segment displays at a rate of 1 per second for 3 minutes. Participants are asked to press the space bar whenever the segment forms a number during the first half of the test or a letter during the latter half. Participants are shown 180 items, 60 of which are target stimuli. Intra-individual variability in reaction time was calculated as the standard deviation in reaction time for true positive responses. RTV was log transformed to an approximately normal distribution, and the natural log of RTV was used in further analysis.

##### 1.1.2. **The Thematically Organised Psychosis Research Study (TOP)**

The TOP study is a case-control study that recruited participants of European ancestry, born in Norway, from the Oslo region.^10^ Cases were individuals with a diagnosis of schizophrenia or bipolar disorder. Diagnosis was confirmed using the Structured Clinical Interview for DSM-IV-TR-axis I disorders.^2^ Healthy controls were randomly selected from statistic records of individuals in the same catchment area as cases. Informed consent was provided by all participants and the human subjects protocol was approved by the Norwegian Scientific-Ethical Committee and the Norwegian Data Protection Agency.

DNA was extracted from blood and saliva samples collected at enrolment. Genotyping was performed using the Human Omni Express-24 v.1.1 (Illumina Inc., San Diego, CA, USA) at deCODE Genetics (Reykjavik, Iceland). Pre-imputation quality control was performed using PLINK 1.9^3^ and involved removal of SNPs with genotyping rate < 95%, Hardy-Weinberg disequilibrium test p-value < 10-4, high rate of Mendel errors in eventual trios or significant (False Discovery Rate < 0.5) batch effects. Whole individual genotypes were excluded if they had low coverage (< 80%) or high likelihood of contamination (heterozygosity > 5 standard deviations above the mean). The quality-controlled genotypes were phased using Eagle^5^, and missing variants were imputed with MaCH^11,12^ using version 1.1 of the trans-ethnic reference sample put together by the haplotype reference consortium (HRC)^4^. High quality variant sets from the quality control procedure were selected to compute individual’s genetic principal components representing loadings along the 20 first eigenvectors of the pairwise genetic covariance matrix of a sub-sample of unrelated individuals from the HRC panel. Following the quality control and imputation procedure, variants with information score < 0.8 or minor allele frequency lower than 0.01 were removed. In addition, individual genotypes imputed with < 75% confidence were set to missing, the remaining ones were converted to best guess hard allelic dosages.

A subsample of participants completed a battery of cognitive tests, including the continuous performance test – identical pairs (CPT-IP) version. The CPT-IP is a measure of sustained attention and during the test, participants are asked to press the mouse key as quickly as possible when two identical pairs of numbers are presented in sequence.^13^ The CPT-IP included a 2-digit, 3-digit, and 4-digit target condition. Participants completed 150 trials for each target condition, 30% of the trials were target trials and required a response. For this study, reaction time variability was calculated as the standard deviation in response time across trials with correct responses for the 2-digit target condition of the task.

#### 1.2. **Supplementary Methods**

##### **1.2.1. Phenotype Association Analyses**

Association analyses were conducted between the 17 selected phenotypes and RTV using linear regression. Each trait was used as the independent variable in a linear regression with RTV as the dependent variable. Z-score standardization was applied to continuous phenotypes and the rank-based Inverse Normal Transformation was applied to RTV prior to conducting the association analysis. Age, sex, scanner site, and Euler number were included as covariates in the regression models for the 10 imaging measures. For the 7 subcortical volumes, total intracranial volume was included as an additional covariate. Age and sex were included as covariates in the association analyses for educational attainment, general cognitive ability, neuroticism, Alzheimer’s disease, attention deficit hyperactivity disorder, posttraumatic stress disorder and schizophrenia. Bonferroni correction for multiple testing was applied and the threshold for significance was p < 2.94 x 10^-8^.

### 2. **Supplementary Figures**

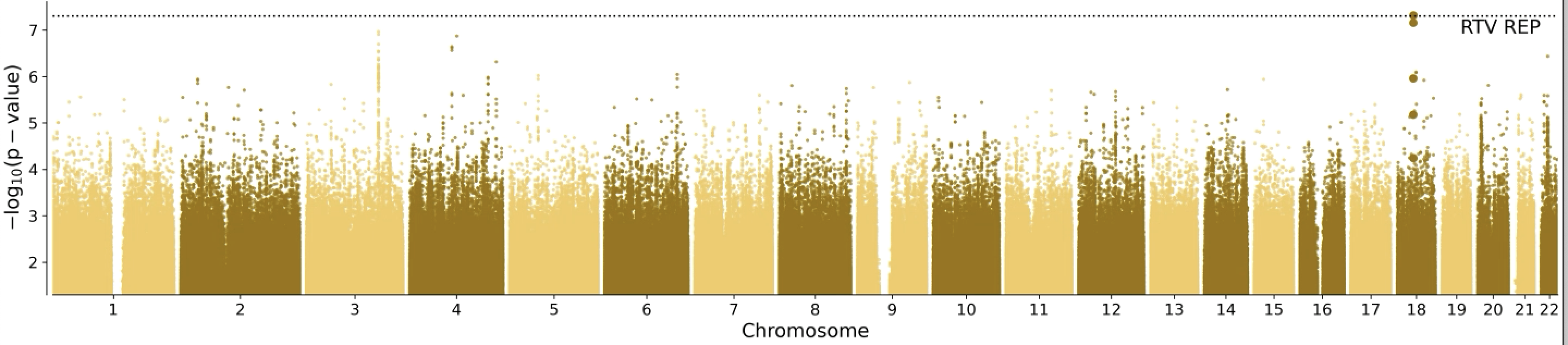

A

B

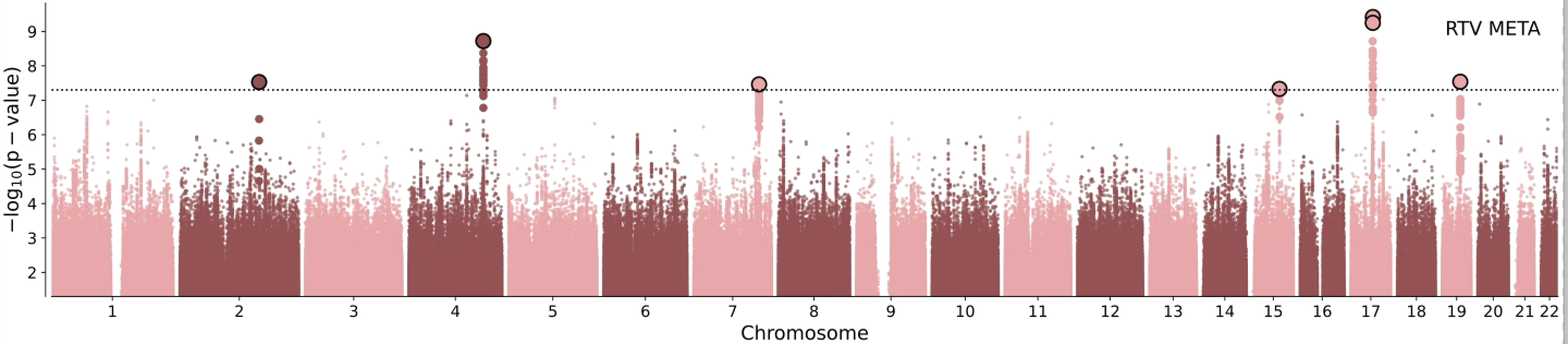

*PSMD14*

*GATB*

*EXOC4*

*TBC1D21*

*LRRC37A2*

*ZNF567*

**Supplementary Figure 1.** Manhattan plots of the results for the replication GWAS (**A**) and discovery and replication GWAS meta-analysis (**B**). The Manhattan plot shows the -log10 transformed two-tailed *P*-values of SNP associations with RTV in a linear regression model against their chromosomal position. The dotted line indicates a genome-wide significance threshold of 5 x 10^-8^. The lead SNPs from the GWAS are outlined in black and the candidate SNPs are shown in bold.

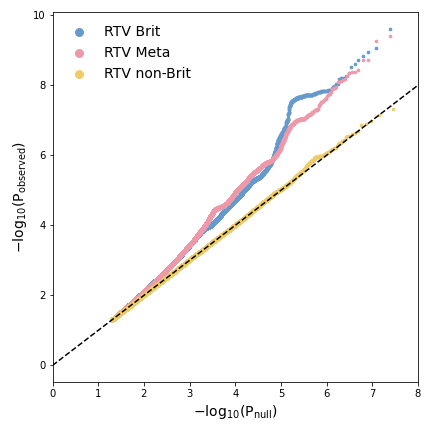

Discovery

Meta-analysis

Replication

**Supplementary Figure 2. .** Quantile-quantile plot of expected under null (no association, x axis) versus observed (y axis) -log_10_ p-values for the discovery (blue), replication (pink), and discovery and replication (yellow) GWAS meta-analysis.

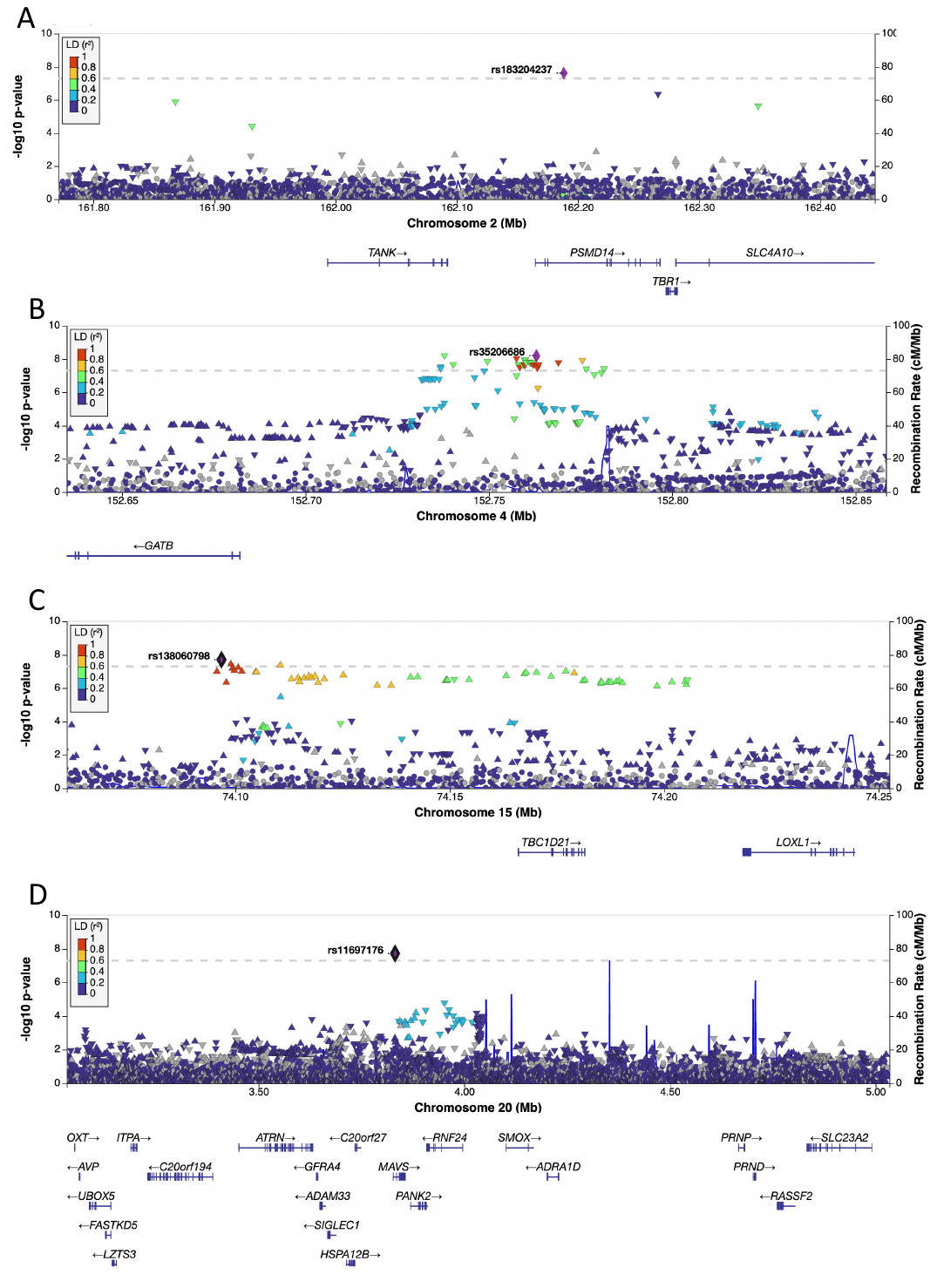

**Supplementary Figure 3.** Regional plots for rs183204237 (**A**), rs35206686 (**B)**, rs138060798 (**C**), and rs11697176 (**D**). The dotted line denotes a genome-wide significance threshold of 5 × 10^–8^. SNPs in the genomic risk loci are colour-coded as a function of their linkage disequilibrium r^2^ to the lead SNP in the region.

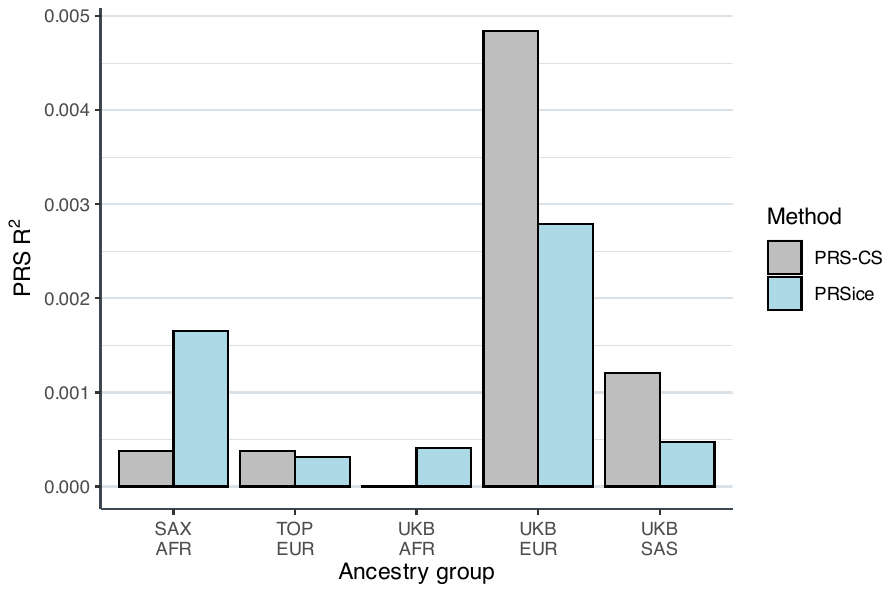

***

***

***

*

**Supplementary Figure 4.** Prediction of RTV by polygenic score (PGS) in 5 independent cohorts. The predictive accuracy of the PGS (R^2^) was assessed in each cohort for a PGS calculated using two methodologies, PRSice and PRS-CS. PRSice PGS were calculated using all single nucleotide polymorphisms surviving LD pruning from the discovery GWAS (*p-value* threshold of 1).

**p*<0.05, ****p*<2.63 x 10^-3^
